# CovidRiskCalc: An online app to calculate the risk of COVID infection in a gathering

**DOI:** 10.1101/2020.12.01.20241646

**Authors:** Marc Artzrouni

## Abstract

CovidRiskCalc is an evidence based online app which calculates the risk of COVID-19 infection for a person coming into contact during a specific event/gathering with a group of individuals, some of whom may be infected (available at CovidRiskCalc.eu). The user is helped in providing a rough estimate of the COVID-19 prevalence rate in the group. She also inputs the size of the group, the number (and duration) of her contacts and the level of precautions (masks, social distancing, etc.). The app calculates the user’s risk of transmission in a single infected contact; her probability of infection during the entire event and the number of new infections within the group. Two numerical examples are given. The tool, designed for both professionals and the general public, thus quantifies the risks of infection in special populations (social gatherings, prisons, etc.), but also in general ones (stores, stadiums, etc.).

## Introduction

As 2020 comes to a close the world will have surpassed 70 million reported cases and at least 1.5 million deaths related to COVID-19. Still, with vaccination campaigns now underway in the UK, US, China, Russia and elsewhere, the end of the tunnel is in sight. In the meantime, governments struggle with “anti-contagion” policies made with little information on the best ways to limit the spread of a disease that is transmitted mostly through close personal contact [1, 2]. These policies are often met with resistance because people have difficulty assessing their individual risk of infection. In the same way they have an intuitive grasp of the risk they take when they go bungee jumping or drive a car, people can make better decisions if they know at least *orders of magnitudes* for the risk of becoming infected with `SARS-CoV-2` during a subway ride, in a family gathering, or while shopping in a supermarket.

A number of online COVID-19 risk calculators have been developed to shed light on these risks. Brown University’s Center for Digital Health has developed [MyCOVIDRisk](https://mycovidrisk.app/) which provides non-numerical “low-medium-high” estimates of your risk based on the particular type of activity you engage in. Another calculator estimates probabilities of at least one person being infected in a group [4]. In both these cases the COVID prevalence needed to estimate the risks is “geolocalized” on the basis of a US address (or county) provided by the user. We complement these efforts here with “CovidRiskCalc”, a bilingual (English/French) online app which is designed for general use and can be applied to any population (available at CovidRiskCalc.eu).

## Method

### i. Probabilistic model

The app is based on a model which quantifies a user’s risk of infection during an event/gathering that brings together a group of individuals characterized by a COVID-19 prevalence *p* that is specified, with help, by the user. An “event” is defined by the size *n* of the group; the number *m* of contacts you will have within the group; the duration *dc* of each contact and one of three possible levels of precautions (i.e. the extent of mask wearing, physical distancing, hand hygiene, etc.). These low, average, and high levels are defined through three decreasing probabilities of transmission *ptr* modeled as functions of *dc* on the basis of the COVID-19 outbreak at a choir practice in Washington State early in the epidemic [4]. (see MODEL tab in the app for details).

The model assumes random movements and contacts formulated as a metaphorical “urn problem”: the user draws one after the other (with replacement) the specified number *m* of contacts among the *n* members of the group, a random number of which are infected (binomial distribution - see MODEL tab). The app calculates the following three metrics:

i. The user’s probability of transmission for each *infected contact* in the group (*ptr*).
ii. The user’s (“takeaway”) *overall probability*^*1*^ of getting infected during the event (*P*).
iii. The number of new infections if all members of the group also mingle with *m* random contacts each of duration *dc*. (*E*).

In the context of iii a byproduct of the results is the event-specific reproduction number R_0_, i.e. the average number of new infections generated by an infected person following her *m* contacts. An average *m(1-p)* of these contacts are with susceptible members of the group and therefore R_0_ = *m(1-p)ptr*.

### ii. Prevalence input

The assumed prevalence *p* will depend on the event. You may hypothesize a 5 or 10% prevalence level in a highly exposed *special population* such as a prison or meat packing plant. On the other hand you will need a (lower) *population-level* prevalence if you are in a supermarket or a stadium.

The fact that the infectious period lasts roughly 10 days means that a *percentage prevalence p* can be approximated as 1/100^th^ the *daily incidence rate* per 100,000 prevailing in the area - a statistic that is often available at local and national levels (See MODEL tab). However, the reported daily incidence rate suffers from “ascertainment bias”, i.e. because of a lack of testing it underestimates the true incidence by a factor of 5 or 10, although this bias has decreased over time [3]. Multiplying the daily incidence rate per 100,000 by such a factor before dividing by 100 thus provides a rough estimate of *p*. In order to assess the sensitivity of the results to *p* the app provides outputs for a range of plausible values of *p*, in addition to the one specified by the user.

## Applications

### i. White House superspreader event

On Sept 26, 2020 approximately *n*=150 guests attended a White House reception to announce the nomination of Judge Amy Coney Barrett to the Supreme Court. There were many hugs and handshakes but few mask [4]. Our data point is the seven people who are presumed to have become infected during the event. With the goal of replicating this outcome with plausible parameter values, we assume a prevalence *p*=2.9% and a hypothetical uninfected guest who mingled with no precautions for a total of 120 minutes, spending *dc*=12 minutes with each one of *m*=10 random attendees (Figure 1; these parameter values are the default ones when opening the app). The calculated probability of transmission *ptr*=16.87% is significantly larger than the overall (“takeaway”) probability *P* = 4.76%. This can happen because *P* is an average assessment of the risk that accounts for the fact that the user may not have any infected contact. The predicted average number of new infections is *E* = 6.89, in line with the seven new infections reported after the event. The reproduction number is R_0 =_ *m(1-p)ptr* = 1.64. The sensitivity analysis on *p* (Figure 1) shows that *E* is 7.12 if *p*=3% and 1.24 if *p*=0.5%.

**Figure 1.**
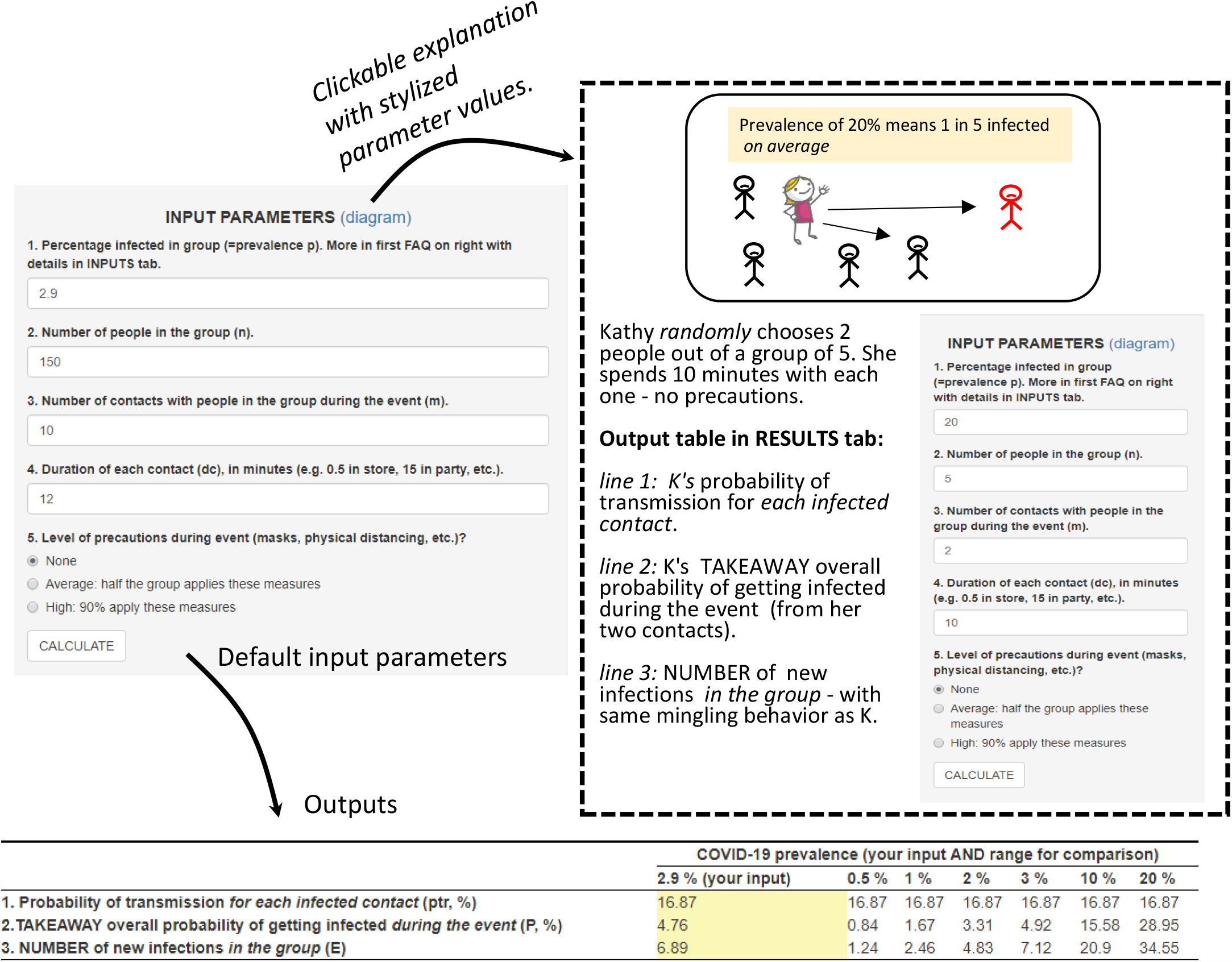
Screenshot of CovidRiskCalc app with default inputs and outputs for illustrative White House Sept 26 superspreader event.

### ii. London store visit

On December 12 Greater London reported a 7-day average daily incidence of 41.2 cases per 100,000. Upon dividing by 100 and multiplying by a correction factor of 5 we obtain an estimated prevalence of *p*=41.2/100×5 = 2.06%. We assume you enter a London store with 50 people in it. You spend two minutes interacting with each one of 10 workers/customers while observing an average level of precautions. The app calculates that your chance of contracting the disease during one infected contact is *ptr* =1.53%, for an overall chance of infection *P* = 0.31%. The reproduction number is *R*_*0*_ = 0.15; *E* = 0.15 new infections meaning that the same event taking place in 100 different stores would result in an average 15 infections. (Up to rounding errors *R*_*0*_ and *E* are equal in this example because the expected number of infected people in the gathering is 50 ×2.06% ≈ 1.)

## Discussion

CovidRiskCalc can be used by professionals and the general public alike to assess the risk of infection during an event/gathering in any setting or population, be it the workplace, a family gathering, prison or stadium^2^. It relies critically on an estimated prevalence that is poorly known - hence the sensitivity analysis. The only thing that needs changing in order to use the tool for another infectious disease is the model for the probability of transmission *ptr*.

The app has limitations due to the model’s simplifying assumptions, namely the uniform mixing and person-to-person transmission, which preclude a spread of the disease via surface contacts or travelling aerosol droplets. Given these assumptions and the uncertainties concerning both the prevalence *p* and the probability of transmission *ptr*, the results should be viewed as orders of magnitudes only – although the model has been validated at least for the well-documented event at the White House on Sept 26.

When driving to a social event or a stadium during the COVID-19 pandemic, you not only have an intuitive grasp of your chance of a flat tire - you can now have some idea of your risk of infection and of the approximate number of new cases during the event.

## Data Availability

data entered by user

https://CovidRiskCalc.eu

## Footnotes

1 This “takeaway” result is highlighted in the app for the benefit of the casual, non-professional user.

2 When navigating to CovidRiskCalc.eu on a mobile phone you are automatically taken to “CovidRisk Lite”, a compact, simplified version of the app designed for the public. It offers the user a choice of only four levels of prevalences.

